# Visual motion and decision-making in dyslexia: Evidence of reduced accumulation of sensory evidence and related neural dynamics

**DOI:** 10.1101/2021.05.26.21257878

**Authors:** Catherine Manning, Cameron D. Hassall, T. Hunt Laurence, Anthony M. Norcia, Eric-Jan Wagenmakers, Margaret J. Snowling, Gaia Scerif, Nathan J. Evans

**Affiliations:** Department of Experimental Psychology, University of Oxford, UK; School of Psychology and Clinical Language Sciences, University of Reading, UK; Department of Psychiatry, University of Oxford, UK; Department of Psychology, Stanford University, USA; Faculty of Social and Behavioural Sciences, University of Amsterdam, The Netherlands; School of Psychology, University of Queensland, Australia

## Abstract

Children with and without dyslexia differ in their behavioural responses to visual information, particularly when required to pool dynamic signals over space and time. Importantly, multiple processes contribute to behavioural responses. Here we investigated which processing stages are affected in children with dyslexia when performing visual motion processing tasks, by combining two methods that are sensitive to the dynamic processes leading to responses. We used a diffusion model which decomposes response time and accuracy into distinct cognitive constructs, and high-density EEG. 50 children with dyslexia and 50 typically developing children aged 6 to 14 years judged the direction of motion as quickly and accurately as possible in two global motion tasks, which varied in their requirements for segregating signal-from-noise. Following our pre-registered analyses, we fitted hierarchical Bayesian diffusion models to the data, blinded to group membership. Unblinding revealed reduced evidence accumulation in children with dyslexia compared to typical children for both tasks. We also identified a response-locked EEG component which was maximal over centro-parietal electrodes which indicated a neural correlate of reduced drift-rate in dyslexia, thereby linking brain and behaviour. We suggest that children with dyslexia are slower to extract sensory evidence from global motion displays, regardless of whether they are required to segregate signal-from-noise, thus furthering our understanding of atypical perceptual decision-making processes in dyslexia.

## Introduction

There has been longstanding interest in the role of visual processing in relation to the reading difficulties experienced by people with developmental dyslexia (e.g., Hinshelwood, 1896; Lovegrove et al., 1980; Morgan, 1896; Orton, 1937). One important aspect of visual processing that appears to develop atypically in those with dyslexia is the ability to process visual motion information. This ability is important for everyday functioning as it allows people to interact with a dynamic world, as well as feeding into a range of functions such as scene segmentation, depth perception and object recognition (Braddick et al., 2003).

Difficulties in global motion processing, demonstrated using tasks that require integrating visual information over space and time, have been widely reported in dyslexic individuals relative to those without dyslexia (Benassi et al., 2010). The most commonly used task is the motion coherence task, in which participants are asked to detect or discriminate motion carried by coherently moving signal dots in a field of randomly moving noise dots (Newsome & Paré, 1988). In this task, dyslexic individuals tend to have higher psychophysical thresholds than typical individuals – that is, they require more dots to be moving coherently in order to perform at the same level of accuracy as those without dyslexia (Cornelissen et al., 1995; Hansen et al., 2001; Pellicano & Gibson, 2008; see Benassi et al. 2010, for meta-analysis). Relationships between reading ability and global motion sensitivity have also been reported in individuals within the normal range of reading ability (Egset et al., 2020; Johnston et al., 2016; Witton et al., 1998, but see also Edwards & Schatschneider, 2019). Yet the nature of the relationship is still being debated, with some researchers proposing a causal relationship between motion sensitivity and reading ability (Boets et al., 2011; Gori et al., 2016) while others challenge this notion (Goswami, 2015; Joo et al., 2017; Olulade et al., 2013; Piotrowska & Willis, 2019).

Perhaps the most influential explanation of atypical global motion processing in dyslexia is reduced sensitivity to rapid temporal information originating from a deficiency in the magnocellular system (Livingstone et al., 1991; Stein, 2001, 2019; Stein & Walsh, 1997) or the related dorsal stream (Braddick et al., 2003; Hansen et al., 2001). The visual system is broadly divided into two processing streams, which are first distinguished at the retina and project to separate layers in the lateral geniculate nucleus and onto superficial layers of the striate cortex (Livingstone & Hubel, 1988; Tootell et al., 1988). Compared to the parallel parvocellular-ventral stream, the magnocellular-dorsal system has cells with properties that make it particularly specialised for motion and depth perception (Livingstone & Hubel, 1988). Although motion coherence tasks are often described as tests of magnocellular processing (e.g., Ebrahimi et al., 2019; Gori et al., 2016), elevated motion coherence thresholds do not necessarily originate from differences in the magnocellular system (Skottun, 2011; Skottun & Skoyles, 2006, 2008; Skottun, 2016). Alternative accounts suggest that dyslexic individuals have difficulty filtering out the motion of the randomly moving noise dots in coherent motion tasks (“noise exclusion”; Conlon et al., 2012; Sperling et al., 2006; see also Ziegler et al., 2009) or difficulties integrating over space and time (Benassi et al., 2010; Hill & Raymond, 2002; Raymond & Sorensen, 1998).

Yet, despite their detailed focus on the sensory characteristics of visual motion stimuli, these accounts have given little consideration to the dynamic processes leading to atypical behavioural responses in dyslexia, and in particular, have not considered whether decision-making processes are affected. A recent EEG study by Toffoli et al. (under revision) reported that activity evoked by coherent motion was similar in children with and without dyslexia at early timepoints, but that group differences emerged at later timepoints, at around 430 ms after the stimulus (see also Schulte-Körne et al., 2004), and differences were also apparent preceding and around the time of the response. These findings suggested that, rather than children with dyslexia differing in early sensory encoding stages of motion processing (cf. the magnocellular hypothesis, Stein 2001), children with dyslexia might instead differ from typically developing children in later processing stages which could reflect sustained visual responses, decision-making, metacognitive processes and/or response generation.

Building on this suggestion, here we explicitly modelled the decision-making process using a popular cognitive model of accuracy and response time: the diffusion model (Stone, 1960; Ratcliff, 1978). This framework allows us to identify the locus of atypical processing in dyslexia, by determining whether children with dyslexia are limited by decision-related or non-decision-related processes. The decision is modelled as a noisy evidence accumulation process from a starting point towards one of two decision bounds (e.g., “left” or “right” decision bounds for a motion discrimination task; see Figure 1 and Evans & Wagenmakers, 2020, for review). The main parameters of the model are drift-rate (reflecting the rate of evidence accumulation), boundary separation (reflecting response caution), starting point (reflecting bias towards one of the response options) and non-decision time (reflecting sensory encoding and response generation). This modelling approach will not only help uncover the reasons for atypical behavioural responses to visual motion information in dyslexia, but also offers two further advantages. First, the resulting parameters may be more sensitive to group differences than accuracy or response time measures alone (Stafford et al., 2020) and second, the parameters relate well to neural measures (Kelly & O’Connell, 2013; Manning et al., 2021; Turner et al., 2015). Accordingly, we combined our diffusion model analysis with a neural measure sensitive to the dynamic processes contributing to behavioural responses (EEG), allowing us to link levels of explanation from brain to behaviour.

**Figure 1.**
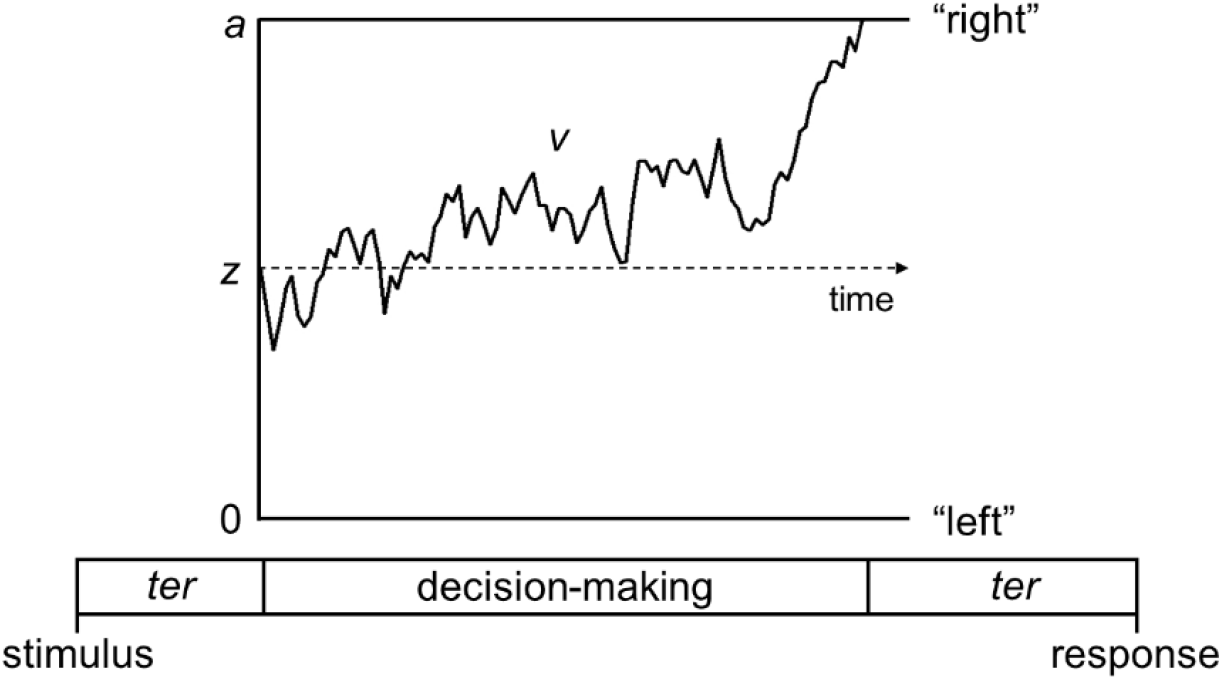
Schematic representation of the decision-making process in the diffusion model for a trial with rightward motion. Decision-making process represented as a noisy accumulation of evidence from a starting point, *z*, towards one of two decision bounds. In our motion tasks, the decision bounds correspond to left and right responses. Boundary separation, *a*, represents the width between the two bounds and reflects response caution. Drift-rate, *v*, reflects the rate of evidence accumulation. Non-decision time, *ter*, is the time taken for sensory encoding processes prior to the decision-making process and response generation processes after a bound is reached.

The diffusion model has been used previously to understand lexical decision performance in individuals with developmental dyslexia and acquired reading impairments (Ratcliff et al., 2004; Zeguers et al., 2011), and has recently been applied to a motion task to investigate relationships with reading ability in children (O’Brien and Yeatman, 2020). In the latter study, a motion coherence task was presented to children with varying levels of reading skill, and it was found that lower reading skill was related to lower drift-rates, wider decision bounds, and more intra-individual variability in both starting point and non-decision time. The reduced drift-rate in poor readers suggests that they accumulate motion evidence more slowly from the coherent motion stimulus than good readers, in line with findings of reduced temporal integration of motion stimuli in individuals with a dyslexia diagnosis (Hill & Raymond, 2002; Raymond & Sorensen, 1998). The wider decision boundaries also suggests that there are differences in decision-making styles between good and poor readers, with poor readers being more cautious in their responses, requiring more evidence to be accumulated before a response is generated.

In this study, we used a diffusion modelling approach to understand which processing stages are affected in children with dyslexia when responding to global motion information. Crucially, we used two different global motion tasks in which participants discriminated overall motion direction, allowing us to better understand the nature of motion processing differences in such children. The first task was a standard motion coherence task, for which elevated thresholds have been widely reported in dyslexia (Benassi et al., 2010). In this task, a proportion of ‘signal’ dots moving left or right were presented amongst randomly moving ‘noise’ dots. The second task was a direction integration task that has not been used previously with dyslexic individuals, whereby dot directions are sampled from a Gaussian distribution, with difficulty being manipulated by varying the standard deviation of the distribution. In this second task, there is no requirement to segregate signal-from-noise. The reason for presenting both tasks to children with dyslexia is to determine whether differences in model parameters are found for both motion tasks, in line with a general motion-processing deficit which may originate from the magnocellular or dorsal stream (Braddick et al., 2003; Stein, 2001), or whether differences in model parameters are found particularly for the motion coherence task, reflecting difficulties segregating signal-from-noise (Conlon et al., 2012; Sperling et al., 2006). When analysing the data we used a blind modelling approach to ensure that modelling decisions were not biased by our hypotheses. We had four primary research questions and hypotheses (see https://osf.io/enkwm for pre-registration):

1. *Do children with dyslexia have reduced drift-rates in a motion coherence task compared to typically developing children?* We hypothesised that children with dyslexia would have reduced drift-rates in the motion coherence task compared to typically developing children, in line with the results of O’Brien and Yeatman (2020) and reports of reduced motion coherence sensitivity in dyslexic individuals (Benassi et al., 2010).
2. *Do children with dyslexia have reduced drift-rates in a direction integration task compared to typically developing children?* If children with dyslexia show difficulties with all global motion tasks (in line with impaired magnocellular/dorsal stream functioning; Braddick et al., 2003; Stein, 2001), then we would expect children with dyslexia to have a reduced drift-rate in this task as well. Instead, if the performance of children with dyslexia in a motion coherence task is limited solely by difficulties with segregating signal-from-noise (Conlon et al., 2012; Sperling et al., 2006), we would expect to see no difference between children with and without dyslexia in this task, as it does not require segregating signal-from-noise.
3. *Do children with dyslexia show increased boundary separation?* We hypothesised that children with dyslexia would have wider boundary separation compared to typically developing children in both tasks, following O’Brien and Yeatman (2020).
4. *Do children with dyslexia show increased non-decision time?* We hypothesised no group differences in overall non-decision time in either task, following O’Brien and Yeatman (2020).

## Methods

### Pre-registration

We pre-registered our inclusion criteria and analysis plan before completing data collection and before commencing analyses (https://osf.io/enkwm).

### Participants

We collected data from 50 children with dyslexia and 60 typically developing children who met our inclusion criteria. Specifically, participants were required to be aged 6 to 14 years (inclusive), have verbal and/or performance IQ scores above 70 (measured using the Wechsler Abbreviated Scales of Intelligence, 2^nd^ edition [WASI-2]; Wechsler, 2011) and to have normal or corrected-to-normal acuity, as measured using a Snellen acuity chart (with binocular acuities of 6/9 or better for children aged 6 to 8 years and 6/6 or better for children aged 9 to 14 years). Children in the dyslexia group were required to have a dyslexia diagnosis (or be in the process of obtaining one, *n* = 1), and to have a reading and spelling composite score of 89 or below, which was computed by averaging the standard scores for the spelling subtest of the Wechsler Individual Achievement Test (WIAT-III; Wechsler, 2017) and the Phonological Decoding Efficiency subtest of the Test of Word Reading Efficiency (TOWRE-2; Torgesen et al., 2012). A cut-off of 89 was chosen to correspond to 1.5 standard deviations below the mean of typically developing children in a similar study (Snowling et al., 2019a, 2019b). Children in the typically developing group were required to have composite scores above 89 and to have no diagnosed developmental conditions. Datasets from an additional 4 typically developing children were excluded due to poor visual acuity (*n* = 1), having a composite score of 89 or below (*n* = 2), or failing to pass criterion on the task (*n* = 1), and datasets from an additional 11 children with dyslexia were excluded due to poor visual acuity (n = 2) or having a composite score above 89 (n = 9).

We then selected 50 typically developing children to best match the children with dyslexia in terms of age and performance IQ using the R MatchIt package (Ho et al., 2011), so that the final dataset included 50 children with dyslexia (24 male) and 50 typically developing children (28 male). As shown in Table 1, the children with dyslexia had slightly higher ages and lower IQ values on average than the typically developing children. EEG data were collected during task performance in 47 typically developing and 44 children with dyslexia (although EEG data were available only in the motion coherence task for one child with dyslexia). The EEG data from these participants were included in a paper investigating responses locked to the onset of coherent motion in typically developing children and children with autism or dyslexia (Toffoli et al., under revision), and the larger group of 60 typically developing children were used to form the comparison group in an autism study (Manning et al., in prep).

**Table 1.** Demographics of participants included in final dataset.

|  | Typically developing<br>(n = 50) | Dyslexia<br>(n = 50) |
| --- | --- | --- |
| Age | 10.65 (2.34) 6.55 – 14.98 | 11.08 (1.87) 7.81 – 14.53 |
| Performance IQ | 109.26 (11.53) 81 – 145 | 99.40 (15.29) 72 – 141 |
| Verbal IQ | 110.60 (8.42) 95 – 127 | 98.56 (10.60) 77 – 118 |
| Full-scale IQ | 111.36 (9.02) 89 – 132 | 98.70 (12.85) 75 – 132 |
| TOWRE-2 PDE | 111.18 (16.53) 81 – 153 | 79.16 (9.45) 51 – 99 |
| WIAT-Spelling | 105.74 (10.21) 80 – 127 | 77.86 (7.96) 58 – 99 |
| Composite score | 108.46 (12.15) 89.5 – 138.0 | 78.51 (7.46) 54.5 – 89.0 |

Note. Data are presented as M (SD) Range.

### Apparatus

The tasks were presented on a Dell Precision M3800 laptop (2048 × 1152 pixels, 60 Hz) using the Psychophysics Toolbox for MATLAB (Brainard, 1997; Kleiner, Brainard & Pelli, 2007; Pelli, 1997). EEG signals were collected using 128-channel Hydrocel Geodesic Sensor Nets connected to Net Amps 300 (Electrical Geodesics Inc., OR, USA) and NetStation 4.5 software. A photodiode attached to the monitor independently verified stimulus presentation timing. Participants used a Cedrus RB-540 response box (Cedrus, CA, USA).

### Stimuli

Stimuli were 100 white, randomly positioned dots (diameter 0.19°) moving at 6°/s within a square aperture (10° x 10°) on a black background, with a limited lifetime of 400 ms. Each trial had a fixation period, a random motion period, a stimulus period, and an offset period, with a red fixation square (0.24° x 0.24°) presented throughout (see Figure 2). By presenting random (incoherent) motion before the stimulus period, we could dissociate evoked responses to directional motion from pattern- and motion-onset evoked potentials. The start of the stimulus period was highlighted to participants with an auditory tone. In the motion coherence task, directional motion (leftward or rightward) was introduced in a proportion of ‘signal’ dots, while the remainder of the dots continued to move in random directions. In the direction integration task, the directions of dots in the stimulus phase were distributed according to a Gaussian distribution with a mean leftward or rightward direction. The fixation period, random motion period and offset period had jittered durations within a fixed range, while the stimulus period was presented until a response or 2500 ms had elapsed. The offset period continued the directional motion to temporally separate motion offset from the response.

**Figure 2.**
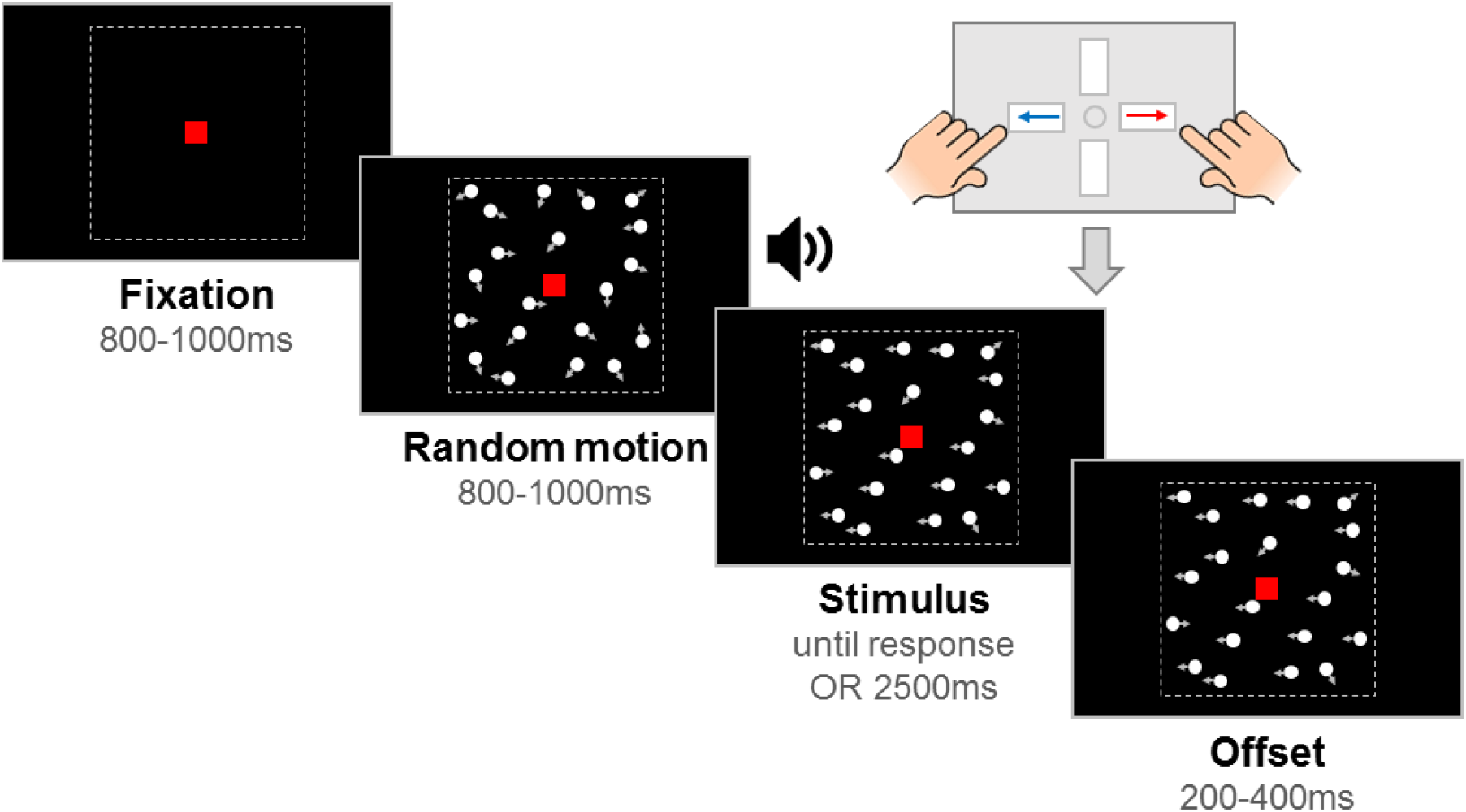
Schematic representation of trial procedure. The trial started with an initial *fixation* period that was followed by a *random motion* period consisting of random, incoherent moving dots, which was in turn followed by a *stimulus* containing leftward or rightward global motion. The child was asked to report the direction using a response box. After the response or after the maximum stimulus duration elapsed (2500 ms), the stimulus remained on the screen for a short *offset* period. Note that arrows (indicating movement) and dotted lines (marking the square stimulus region) are presented for illustration only. The stimulus shown here is from the motion coherence task, where a proportion of dots move coherently. In the direction integration task, dot directions were taken from a Gaussian distribution. Figure reproduced from https://osf.io/wmtpx/ under a CC-BY4.0 license.

### Experimental task procedure

Children completed motion coherence and direction integration tasks within child-friendly games (based on Manning et al., 2019, 2021). Using animations, participants were told that fireflies were escaping from their viewing boxes, and they were asked to tell the zookeeper which way the fireflies were escaping. There were 10 ‘levels’ of the game. Levels 1-5 corresponded to one task (either motion coherence or direction integration), and Levels 6-10 corresponded to the other task, with the order of tasks being counterbalanced across participants. Levels 1 and 6 were practice phases, and the remaining 4 levels for each task were experimental blocks. In the motion coherence task, difficulty was manipulated by varying the proportion of coherently moving dots, and in the direction integration task, difficulty was manipulated by varying the standard deviation of the Gaussian distribution from which the dot directions were sampled.

In the practice phases, four demonstration trials were presented with no random motion phase and an unlimited stimulus phase, so that the experimenter could explain the task. Participants reported stimulus direction using a response box. The first two demonstration trials were ‘easy’ (100% coherence or 1° standard deviation), and the last two were more difficult (75% and 50% coherence, or 10° and 25° standard deviations). Following the demonstration trials, there were up to 20 criterion trials with a coherence of 95% or a standard deviation of 5°. These trials introduced the random motion phase. It was explained to participants that the fireflies would be going “all over the place” at first, and that they must wait for an alarm (auditory beep) before deciding which way the fireflies were escaping. A time limit was enforced, with visual feedback presented on the screen if participants did not respond within 2500 ms (“Timeout! Try to be quicker next time!”). Feedback on accuracy was given for responses made within the time limit (“That was correct!”, or “It was the other way that time”). When participants met a criterion of four consecutive correct responses, no more criterion trials were presented. Next, there were eight practice trials of increasing difficulty (motion coherence task: 80%, 70%, 60%, 50%, 40%, 30%, 20%, 10%; direction integration task: 5°, 10°, 15°, 20°, 30°, 40°, 50°, 60°) with feedback as before. Level 1 was repeated for one typically developing child and 2 children with dyslexia who did not meet the criterion of four consecutive correct responses on the first attempt, but passed on the second attempt.

Levels 2-5 and 7-10 each contained 38 trials, with 9 repetitions of each of two difficulty levels (motion coherence task: 30%, 75%; direction integration task: 70°, 30° SD), for each motion direction (leftward, rightward), and an additional 2 catch trials presenting 100% coherent (0° SD) motion. The experimental phase for each task therefore consisted of 152 trials. No trial-by-trial feedback was presented during the experimental phase, apart from a ‘timeout’ message if no response was made within 2500ms after stimulus onset. At the end of each level, participants were given points for their speed and accuracy in the preceding block (computed by (1 / median response time) * the number of correct responses * 2, rounded to the nearest integer). If participants obtained a score under 10, a score of 10 points was given to maintain motivation. Trials were presented automatically, although the experimenter could pause and resume trial presentation if necessary. The experimental code can be found here: https://osf.io/fkjt6/.

### General procedure

The procedure was approved by the Central University Research Ethics Committee at the University of Oxford. Parents provided written informed consent and children gave verbal or written assent. All children took part at the University of Oxford apart from one child with dyslexia who was seen at school without EEG. During the experimental tasks, participants sat 80cm away from the computer screen in a dimly lit room. For children who participated with EEG, we fitted the net prior to the experiment and ensured that electrode impedances were below 50 kΩ. EEG data were acquired at a sampling rate of 500Hz with a vertex reference electrode.

Children were closely monitored by an experimenter sitting beside them. The experimenter provided general encouragement and task reminders, pausing before the start of a trial if needed (e.g., to remind the child to keep still). Children had short breaks at the end of each ‘level’ and a longer break at the end of the first task (at the end of ‘level 5’). During the longer break, electrode impedances were re-assessed for children wearing EEG nets. Children marked their progress through the levels using a stamper on a record card. The children also completed a Snellen acuity test, the WASI-2, the TOWRE-2 and the spelling subtest of the WIAT-III. The whole session took no longer than 2 hours and children were given a gift voucher to thank them for their time.

### Diffusion model analysis

Initially, a blinded analysis was conducted to ensure that modelling decisions were made without being biased by the hypotheses under test. The first author (CM) prepared a blinded dataset in which group membership was randomly permuted (see also Dutilh et al., 2017) and one of the authors (NJE) ran diffusion model analysis on this blinded dataset.

Prior to modelling, trials with response times under 200 ms were removed (corresponding to 0.20% of trials in the typical group and 0.24% of trials in the dyslexia group). Trials without a response (i.e., no response made within the 2500ms deadline) were modelled as non-terminating accumulation trajectories, with the probability of a non-response occurring being the survivor function for the model at the time of the 2500 ms deadline (Evans et al., 2018; Howard et al., 2020; Ulrich & Miller, 1994). These trials accounted for 1.02% of the data in the typical group and 1.26% of the data in the dyslexia group. We fit the data from each task with hierarchical, Bayesian diffusion models with 5 parameters: 1) average drift-rate across difficulty levels *v.mean*, 2) boundary separation *a*, 3) non-decision time *ter*, 4) difference in mean drift-rate between difficulty levels *v.diff*, and 5) starting point *z*. The stochastic noise within the model (*s*) was fixed at 0.1 to solve a scaling problem within the model, as per convention (Ratcliff, 1978). There were 3 hyperparameters for each parameter reflecting the mean (µ) and standard deviation (σ) across the two groups and the difference between groups (δ). Importantly, this parameterization allowed us to explicitly set priors on the differences between groups, which was the key effect of interest within the current study. More specifically, the priors were:

*Data level:*

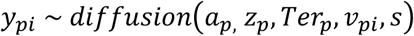

*Parameters:*

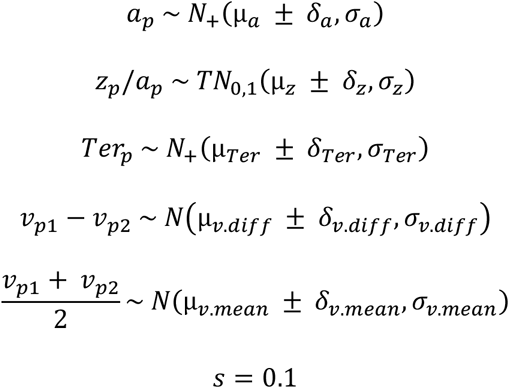

*Hyperparameters:*

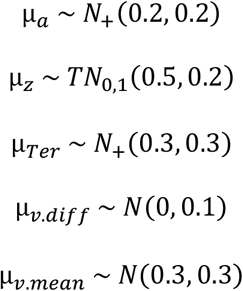

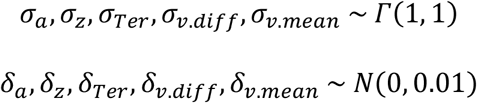

where *y* reflects the data, and subscripts *p* and *i* reflect the participant and difficulty level respectively. The priors for the *µ* and *σ* parameters were based on those used in previous studies implementing hierarchical diffusion models (e.g., Evans & Brown, 2017; Evans & Hawkins, 2019; Evans et al., 2019), and the priors for the δ parameters were based on the “moderately informative priors” used for the differences between conditions in Evans (2019). We used a differential evolution Markov chain Monte Carlo algorithm (DE-MCMC; Ter Braak, 2006; Turner, Sederberg, Brown, & Steyvers, 2013) to sample from the posterior with 15 interacting chains, each with 4000 iterations, the first 1500 of which were discarded as burn-in. We also implemented a migration algorithm (see Turner, Sederberg, Brown, & Steyvers, 2013), where chains were randomly migrated every 14 iterations between iterations 500 and 1100.

As shown in Table 1, the children with dyslexia were on average slightly older and of lower IQ than the typically developing children. As pre-registered, the first author (CM) ran a default Bayesian t-test using the BayesFactor R package (Morey & Rouder, 2018) which revealed weak, inconclusive evidence for the absence of group differences in age (BF in support of group differences = 0.33; Jeffreys, 1961). As we know that diffusion model parameters change with age (Manning et al., 2021), and as we couldn’t conclusively rule out group differences in age, we also ran models which partialled out the effects of age from all of the parameters (using the residuals from the line of best fit between age and each of the parameters), in addition to our standard models. In our pre-registered analysis plan we decided not to control for performance IQ as it may relate to both group membership and decision-making in cognitively relevant ways (Dennis et al., 2009). The analysis files were posted on the Open Science Framework prior to unblinding (https://osf.io/nvwf7/), at which point all models were re-run on the unblinded dataset with correct group membership.

### EEG analysis for joint modelling

We ran exploratory analysis on the unblinded dataset to investigate links between drift-rate and EEG activity. EEG data were band-pass filtered between 0.3 and 40 Hz in NetStation and then exported for further processing in MATLAB using EEGLAB functions (Delorme & Makeig, 2004). We downsampled each participant’s data to 250 Hz and selected only the data between the first fixation onset and the last offset period. We then bandpass-filtered between 0.3 and 40 Hz (due to insufficient attenuation of low frequencies by NetStation filters, Manning et al., 2019) and used EEGLAB’s ‘clean_artifacts’ function to remove bad channels, identify data segments with standard deviations over 15 and correct them using artifact subspace reconstruction (ASR; Chang et al., 2018). Missing channels were then interpolated. We then ran independent components analysis on 3000 ms epochs starting at fixation onset using an Infomax algorithm and subtracted ocular components from the continuous data. Finally, we average re-referenced the data. In line with the behavioural analyses, we excluded triggers for response events made <200 ms or >2500 ms after stimulus onset.

Following previous work, we used a data-driven component decomposition technique to identify spatiotemporally reliable patterns of activity across trials, which has the effect of maximising signal-to-noise ratio (Reliable Components Analysis, Dmochowski et al., 2012; Dmochowski & Norcia, 2015; Manning et al., 2019, 2021). To do this, we epoched each participant’s preprocessed continuous data from -600 ms to 200 ms around each response, and we baselined the data to the last 100 ms of the random motion period. We submitted the baselined epochs for participants in both groups to Reliable Components analysis for each task separately. The forward-model projections of the weights for the most reliable component for each task (which explained 28.7% and 27.1% of the reliability in the motion coherence and direction integration tasks, respectively) are shown in Figure 3. This component resembled the most reliable component found in our previous work (Manning et al., 2021), which in turn resembles the centro-parietal positivity (O’Connell et al., 2012; Kelly and O’Connell, 2013). Build-up of activity in this component has been linked to drift-rate in typically developing children (Manning et al., 2021). To investigate links with drift-rate in the current dataset, we projected each participant’s continuous data through the spatial weights for this component to yield a single component waveform for each participant for each task.

**Figure 3.**
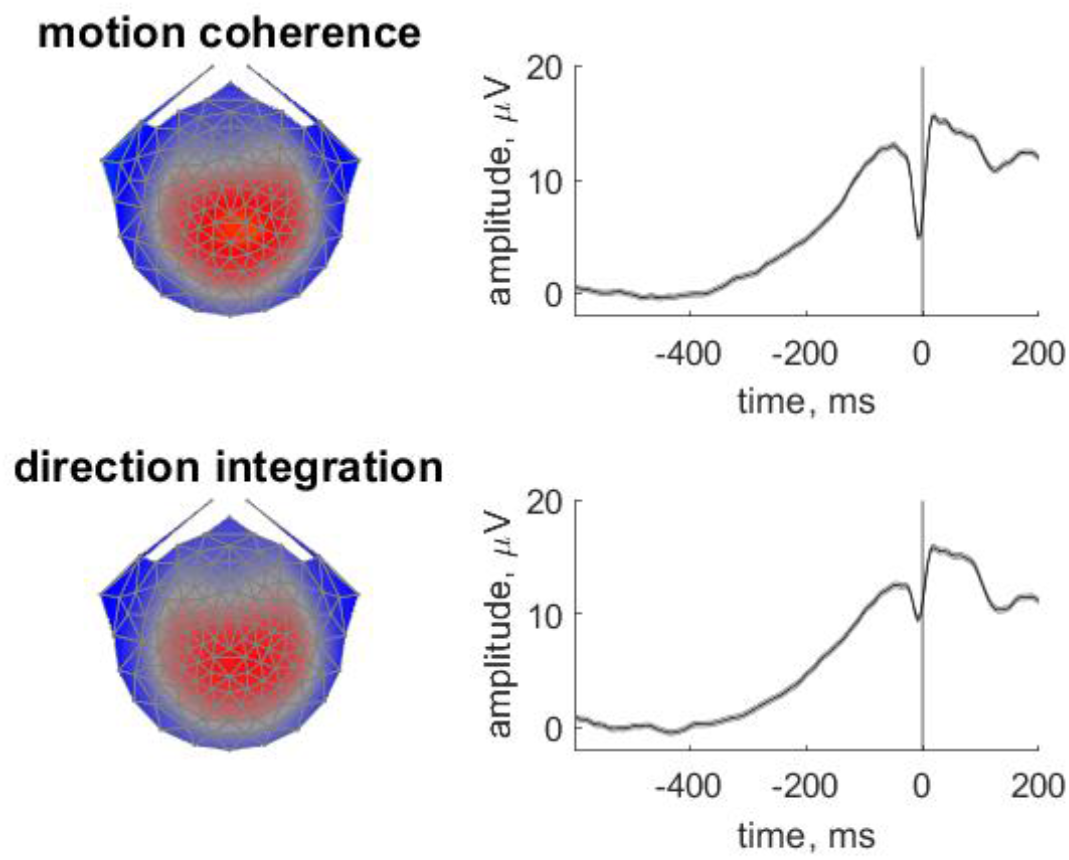
Scalp topographies and temporal dynamics for the most reliable component in the motion coherence and direction integration tasks. Topographic visualisations of the forward-model projections of the most reliable component (left) reflecting the weights given to each electrode following reliable components analysis (RCA) on data from all participants pooled across difficulty level, for the motion coherence task (upper) and direction integration task (lower). The waveforms (right) show the temporal dynamics of the component.

When evaluating group differences in evoked potentials during the decision period, an important consideration is whether the effects are best described as changes in the stimulus-locked or response-locked activity, as these two components will temporally overlap (and the degree of overlap will relate to the subject’s reaction time). Importantly, the extent of overlap could vary between groups and/or conditions (Ehinger & Dimigen, 2019). Thus, to determine the origin of any group differences in evoked potentials, we used a linear deconvolution method to unmix overlapping stimulus-locked and response-locked activity in this component waveform using the Unfold toolbox (Ehinger & Dimigen, 2019). We modelled the continuous waveform for each participant by selecting a time window of -1000 ms to 1000 ms around each stimulus event or response event. We specified a design matrix with predictors for each difficulty level (difficult, easy) for each event type (stimulus, response). We then time-expanded the design matrix by adding a predictor for each timepoint sampled for each event type. We identified segments with amplitudes above ±250 μV using a sliding 2000 ms segment in 100 ms steps, and excluded these segments from the design matrix (mean 2.72% of the data for each participant, range: 0 to 43%). We then fit the deconvolution model resulting in regression weights (betas) for each of the 2 event types, 2 difficulty levels and 500 timepoints, which we used to construct regression waveforms (see Figures 4 and 5).

**Figure 4.**
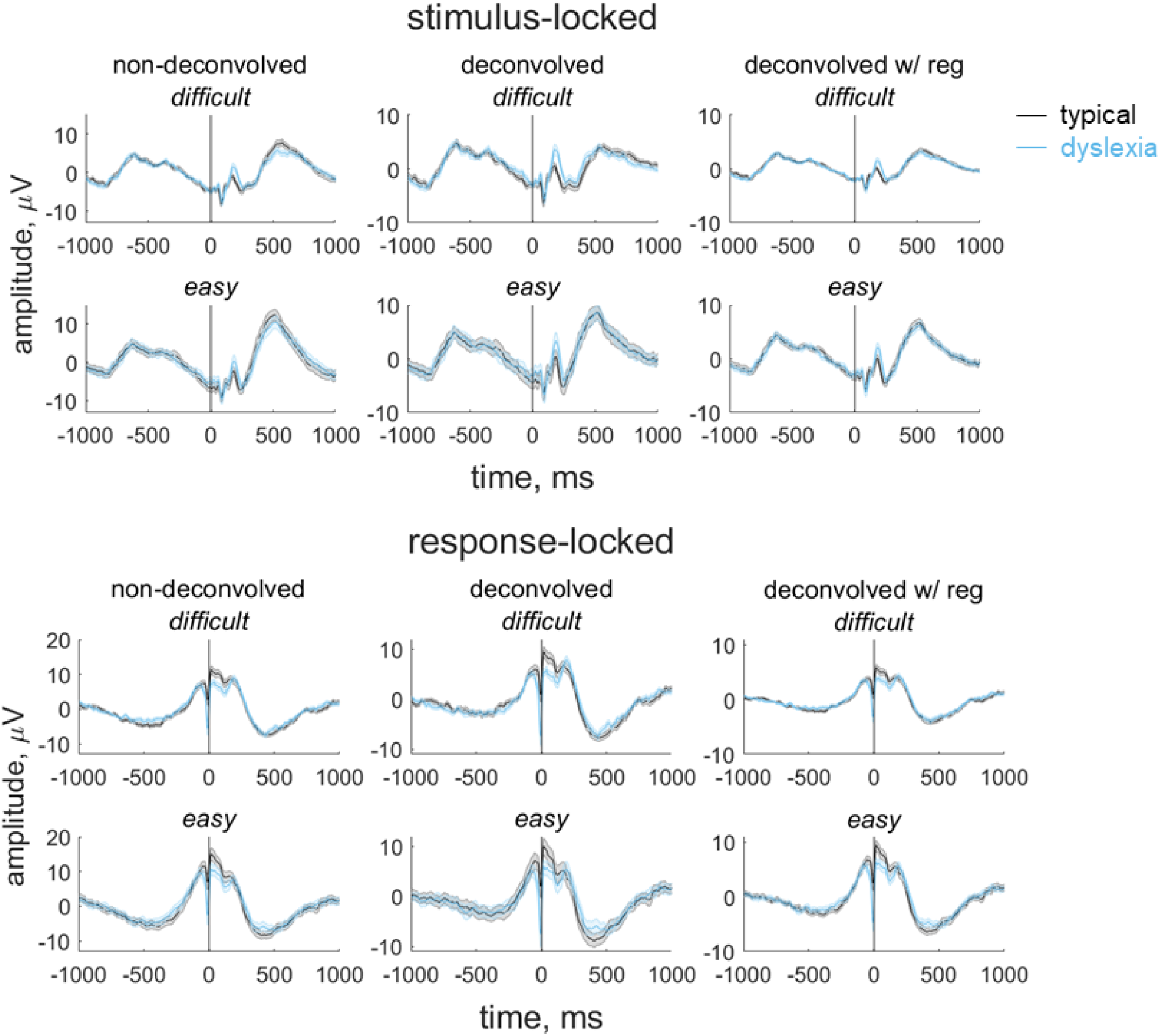
Group average stimulus-locked and response-locked evoked potentials for the motion coherence task. Average (±1SEM) stimulus-locked (upper) and response-locked (lower) evoked potentials for typically developing children (grey) and children with dyslexia (blue) in the motion coherence task for difficult and easy levels. The left column shows non-deconvolved group average waveforms. The central column shows deconvolved group average waveforms (without regularisation). The right column shows deconvolved group average waveforms with regularisation (ridge regression).

**Figure 5.**
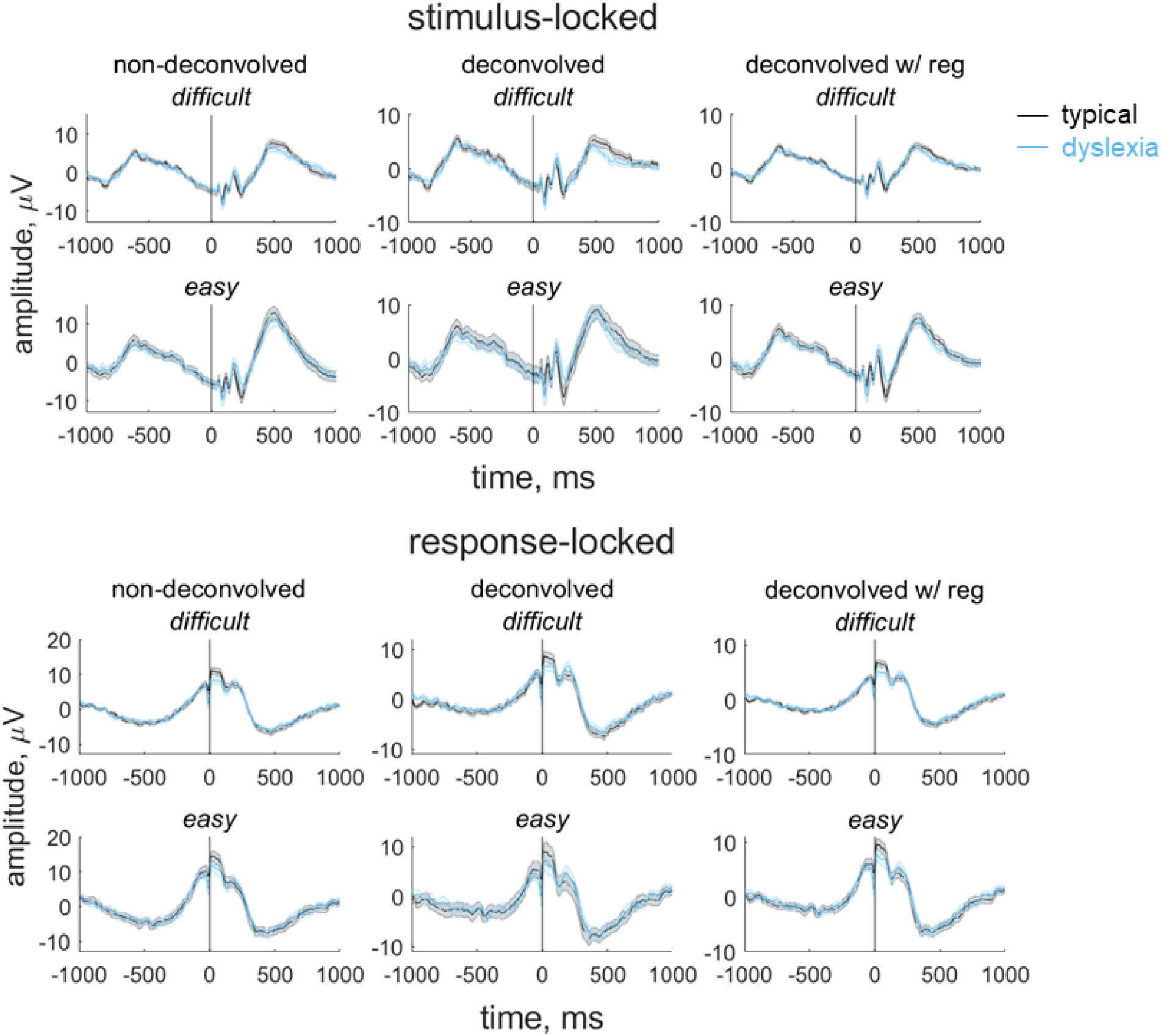
Group average stimulus-locked and response-locked evoked potentials for the direction integration task. Average (±1SEM) stimulus-locked (upper) and response-locked (lower) evoked potentials for typically developing children (grey) and children with dyslexia (blue) in the direction integration task for difficult and easy levels. The left column shows non-deconvolved group average waveforms. The central column shows deconvolved group average waveforms (without regularisation). The right column shows deconvolved group average waveforms with regularisation (ridge regression).

The non-deconvolved waveforms showed amplitude differences between difficult and easy levels (Figures 4 and 5, left column), as to be expected for an EEG measure which reflects the decision-making process. However, these differences across difficulty levels were not evident in the deconvolved waveforms (Figures 4 and 5, central column). The fact that the difference between difficulty levels changed as a result of deconvolution could suggest that the overlap between stimulus- and response-locked activity differs between difficulty levels, due to different RT distributions in each difficulty level. However, we found a difficulty level difference in the non-deconvolved waveforms even when matching the RT distributions for the easy and difficult levels, so that difficulty level differences could not be purely attributed to different RT distributions. We therefore suspected that the beta estimates may be noisy and that the deconvolution technique was overfitting the noise. Therefore, in the final step where we selected EEG measures for inclusion in the diffusion model, we re-ran the deconvolution model using a regularisation method which penalises the squared magnitude of the regression coefficients (ridge regression; see Kristensen et al., 2017) to minimise noise. Using this approach retained the difficulty level differences. Specifically, we found the best regularisation parameter for each participant using cross-validation, and then took the mode across all participants and constrained the regularisation parameter to ensure that differences in regularisation did not contribute to group differences in resulting waveforms. The modal parameter value was 10 for the motion coherence task (5.5 and 10 for the typically developing children and children with dyslexia, separately) and 5 for the direction integration task (5 and 4.5 for the typically developing children and children with dyslexia, separately). We then fit a regression slope to each participant’s average deconvolved waveform for each difficulty level between -200 ms to 0 ms around the time of the response to obtain a slope measure which we entered into the diffusion model and related to drift-rate.

To assess the relationship between drift-rate and the EEG component discussed above, we used a joint modelling approach (Turner et al., 2013, 2015, 2016, Evans et al., 2018; Knowles et al., 2019). Specifically, we estimated additional hyper-parameters for the correlation between the *v.mean* parameter and the average of the EEG measure over difficulty levels *(EEG.mean)*, and between the *v.diff* parameter and the difference in the EEG measure between difficulty levels *(EEG.diff)*. Specifically, this meant that the structure of the original hierarchical model (with age partialled out) was only different for the drift-rate parameter, which was now a bivariate normal with the EEG measure:

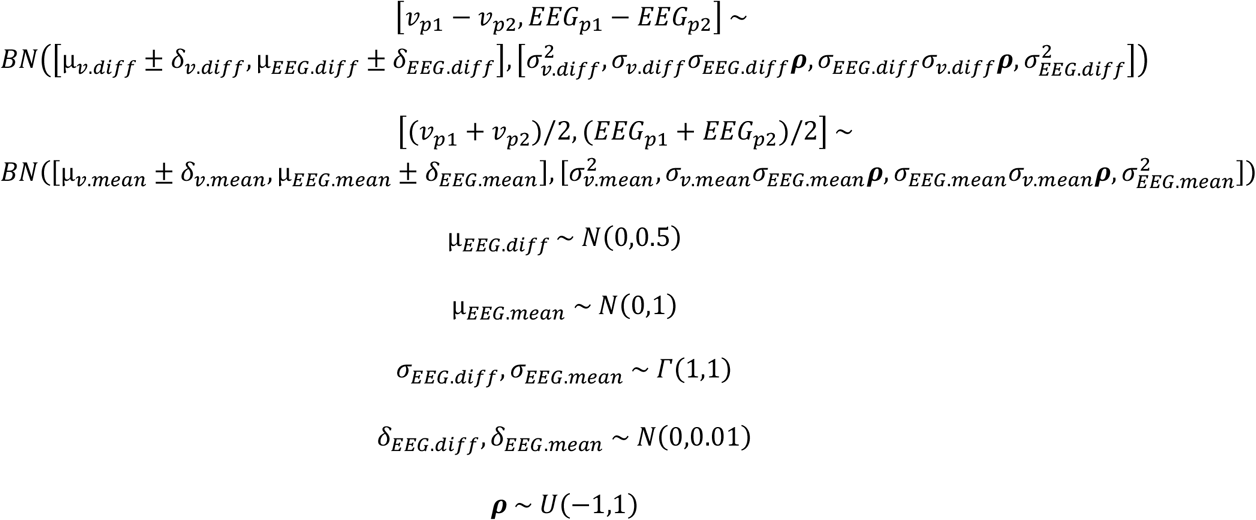

where ***ρ*** refers to the correlation between drift-rate and the EEG measure. Note that we again used DE-MCMC with 15 interacting chains to sample from the posterior of the joint model, though due to the greater computational burden of the model we used 3000 iterations, of which the first 1000 were discarded as burn-in and no migration algorithm was implemented. Furthermore, we estimated two different variants of this joint model: one where the correlations were constrained to be the same across groups, which would allow for the estimation of more precise posteriors due to the limited sample size, and another less constrained version were the correlations were estimated separately for each group.

### Data and code availability

Analysis scripts and output files are available at: https://osf.io/nvwf7/. Data will be madeavailable on the UK Data Service after the manuscript has been accepted for publication.

## Results

### Diffusion modelling of behavioural data

Figure 6 summarises the accuracy and response time data subjected to diffusion modelling. This figure shows that the children with dyslexia had slightly slower median response times compared to typically developing children, overall, and were slightly less accurate in the direction integration task, particularly on the difficult trials. However, there was substantial overlap between the groups with considerable variability within each group. These behavioural data were well-fit by our diffusion models, as shown by the cumulative density functions in Supplementary Figure S1. All chains were well-converged, as reflected by Gelman-Rubin diagnostic values (Gelman & Rubin, 1992) close to 1 (*M* = 1.00, range = 1.00 – 1.07). Figure 7 shows the prior and posterior distributions for the group-level parameters that reflect the difference between groups for each of the 5 parameters (*v.mean, a, ter*, v.diff, *beta*), along with Bayes factors calculated through the Savage-Dickey ratio. Bayes factors above 1 reflect more evidence for the alternative hypothesis of group differences compared to the null hypothesis, whereas Bayes factors below 1 reflect relatively more evidence for the null hypothesis than the alternative hypothesis. We use the heuristic that Bayes factors between 1/3 and 3 constitute only weak, inconclusive evidence (Jeffreys, 1961).

**Figure 6.**
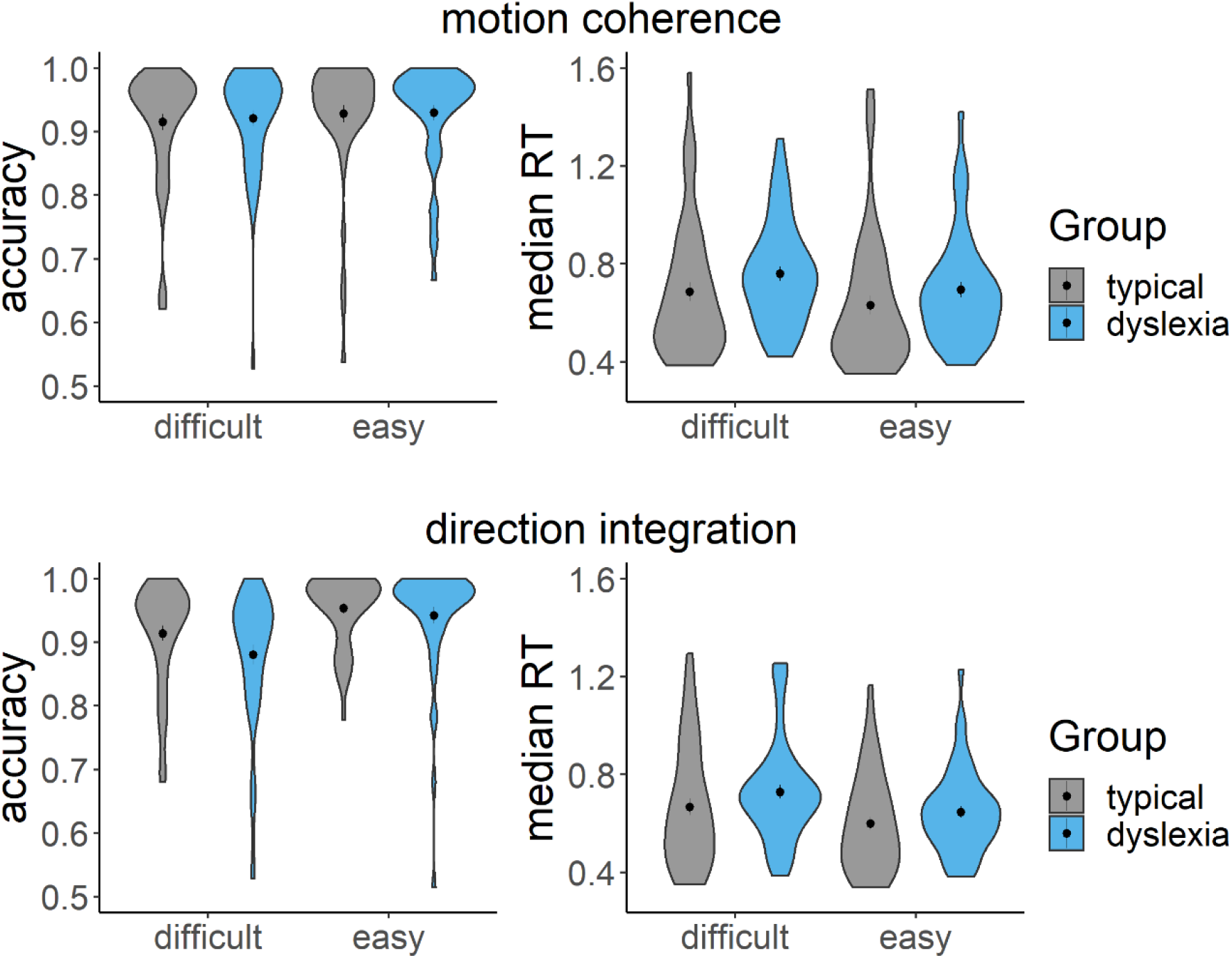
Accuracy and median response time (RT) for correct trials. Violin plots showing the kernel probability density for each group’s accuracy (left) and median RT (s) for correct trials (right) for each difficulty level and each task (upper: motion coherence; lower: direction integration). Data for typically developing children and children with dyslexia are presented in grey and blue, respectively. Dots and vertical lines represent the group mean and ±1 SEM.

**Figure 7.**
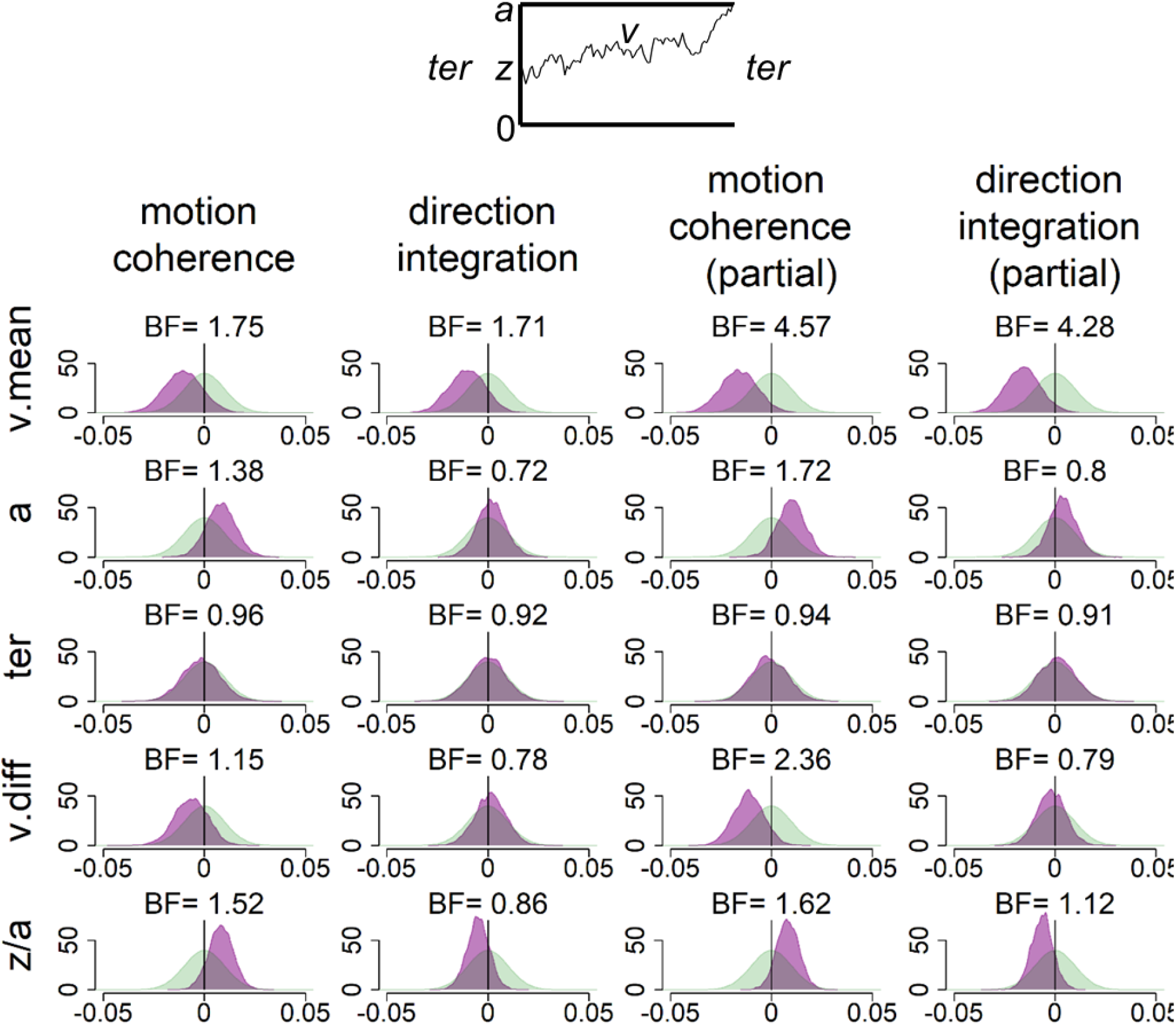
Prior and posterior density distributions. Prior (green) and posterior (purple) density distributions for the group-level parameters reflecting group differences in each of the 5 model parameters (v.mean = mean drift-rate across difficulty levels; a = boundary separation; ter = non-decision time; v.diff = difference in mean drift-rate between difficulty levels; z/a = relative starting point) for each task. The upper inset shows a schematic of the model parameters shown. The leftmost columns show the results of the standard model and the rightmost columns show the results of the model with age partialled out. Negative values reflect lower parameter values in the dyslexia group compared to the typically developing group. BF = Savage-Dickey Bayes factors in favour of the alternative hypothesis (H_1_) over the null hypothesis (H_0_). BF > 1 support H_1._

In support of our first hypothesis, children with dyslexia had reduced drift-rates in the motion coherence task compared to typically developing children, as shown by the leftward shift in the posterior distribution of *v. mean* in Figure 7. When age was partialled out, there was moderate evidence in favour of group differences (BF = 4.57). The evidence was weaker when age was not partialled out (BF = 1.75). Interestingly, the same pattern was found in support of our second hypothesis, with children with dyslexia also showing reduced drift-rates in the direction integration task compared to typically developing children. Again, there was moderate evidence for group differences when age was controlled for (BF = 4.28), but weak evidence when age was not controlled for (BF = 1.71).

Our third hypothesis was that children with dyslexia would show increased boundary separation. Although children with dyslexia did have slightly higher boundary separation compared to typically developing children (indicated by a small rightward shift in the posterior distribution of *a* in Figure 7), particularly in the motion coherence task, the evidence remained inconclusive, even when controlling for age. Our final hypothesis was that there would be no group differences in non-decision time *(ter)* in either task. Figure 7 shows little difference between the groups in this parameter, but the Bayes factors are close to 1, suggesting inconclusive evidence. Therefore, more data would be required to make firm conclusions regarding these hypotheses.

These pre-registered analyses did not control for performance IQ because it could be meaningfully related to both decision-making parameters and group membership, and investigating its contribution to both was beyond the scope of our multi-level modelling approach. However, as there was an indication of a relationship between performance IQ and drift-rate (Figure S2), and as both performance IQ and drift-rate differed between the groups, we investigated these links further with an exploratory analysis which partialled out the effects of both age and performance IQ (Figure S3). In brief, BFs of 2.3 and 2.38 in the two tasks continue to provide weak evidence for group differences in mean drift-rate when both age and PIQ are controlled for.

### Joint modelling of EEG and behavioural data

Figure 8 shows the distribution of slope measures that were extracted from each participant’s deconvolved (with regularisation) response-locked waveform, which were used in joint modelling to explore links between EEG and model parameters, with the effects of age partialled out. As expected, the children with dyslexia had shallower slopes than the typical children, on average, reflecting a more gradual build-up of activity in the centro-parietal component. First we established whether this EEG measure was related to drift-rate across the whole sample, estimating a single correlation for both groups. For both tasks, the EEG measure was positively related to both the mean drift-rate across difficulty levels, though the evidence was only weak in the case of the direction integration task (motion coherence: posterior mean *r* = .44, 95% credible intervals (CI) = [.26, .6], BF = 8869.49; direction integration: posterior mean *r* = .25, CI = [.03, .45], BF = 1.65). The posterior means were in the direction of a positive relationship between the difference in EEG measure and the difference in drift rate between difficulty levels, although the evidence was inconclusive (motion coherence: posterior mean *r* = .22, CI = [-.02, .44], BF = .73; direction integration: posterior mean *r* = .17, CI = [-.08, .4], BF = 0.43; see Figure S2 for scatterplots).

**Figure 8.**
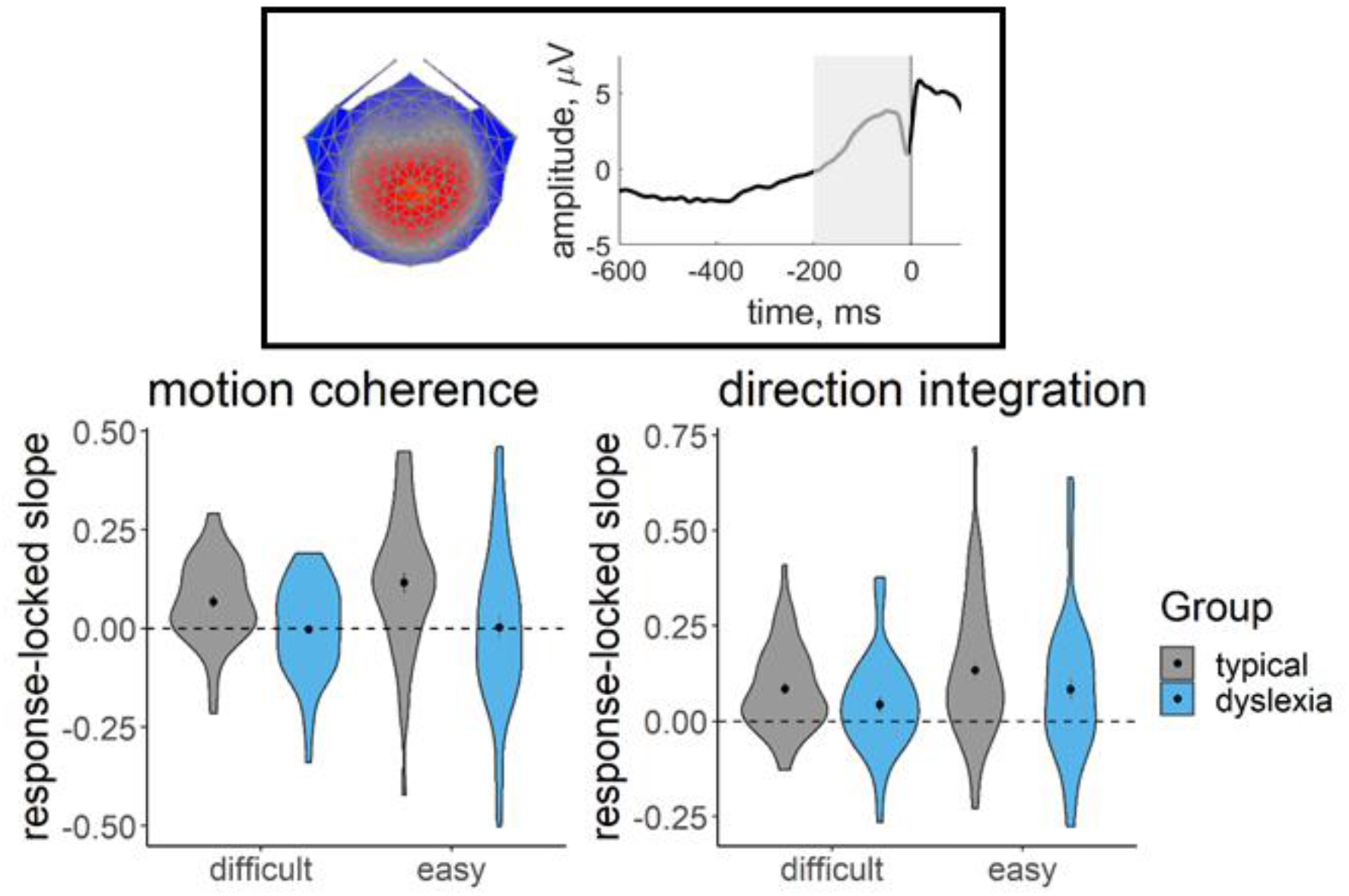
EEG slope measure extracted for inclusion in the joint model. Violin plots showing the kernel probability density for the EEG slope measure extracted for inclusion in the joint model for each group (typically developing: grey; dyslexia: blue) for each difficulty level. The extracted measure was the slope of a linear regression line fitted to each participant’s deconvolved (with regularisation) response-locked waveform, from 200 ms prior to the response to the response (see shaded area of schematic response-locked waveform in inset). The dotted line reflects a flat slope. Dots and vertical lines represent the group mean and ±1 SEM.

Next we fit joint models in which we estimated a separate correlation coefficient between drift-rate and the EEG measure for the children with dyslexia and typical children (Figure 9, Figure S4). Note that our intention was not to explicitly test for differences in correlations between groups, but rather to see if the previous findings seem to hold for each group; any separation between the groups below is intended to merely describe our estimated posterior distributions. A positive correlation can be seen for both groups in the motion coherence task for the mean drift-rate across difficulty levels (typical: posterior mean *r* = .41, CI = [.13, .63], BF = 7.45; dyslexia: posterior mean *r* = .43, CI = [.15, .64], BF = 12.75). The posterior means were in the direction of a positive relationship for the difference in drift-rate between difficulty levels, but the evidence was inconclusive (typical: posterior mean *r* = .18, CI = [-.2, .51], BF = .39; dyslexia: posterior mean *r* = .20, CI = [-.12, .49], BF = .46). The strength of correlations was weaker in the direction integration task, particularly for the typical children, for whom the Bayes factors suggested moderate evidence for no relationship (mean drift-rate across difficulty levels: posterior mean *r* = .10, CI = [-.22, .4], BF = .29; difference between difficulty levels: posterior mean *r* = .04, CI = [-.31, .38], BF = .24). The strength of the correlations in children with dyslexia were slightly stronger than in the typical children, with the mean drift-rate across difficulty levels showing weak evidence for a relationship, though the difference in drift-rate between difficulty levels showed weak evidence for no relationship (mean drift-rate across difficulty levels: posterior mean *r* = .34, CI = [.04, .58], BF = 2.59; difference between difficulty levels: posterior mean *r* = .24, CI = [- .09, .53], BF = .61).

**Figure 9.**
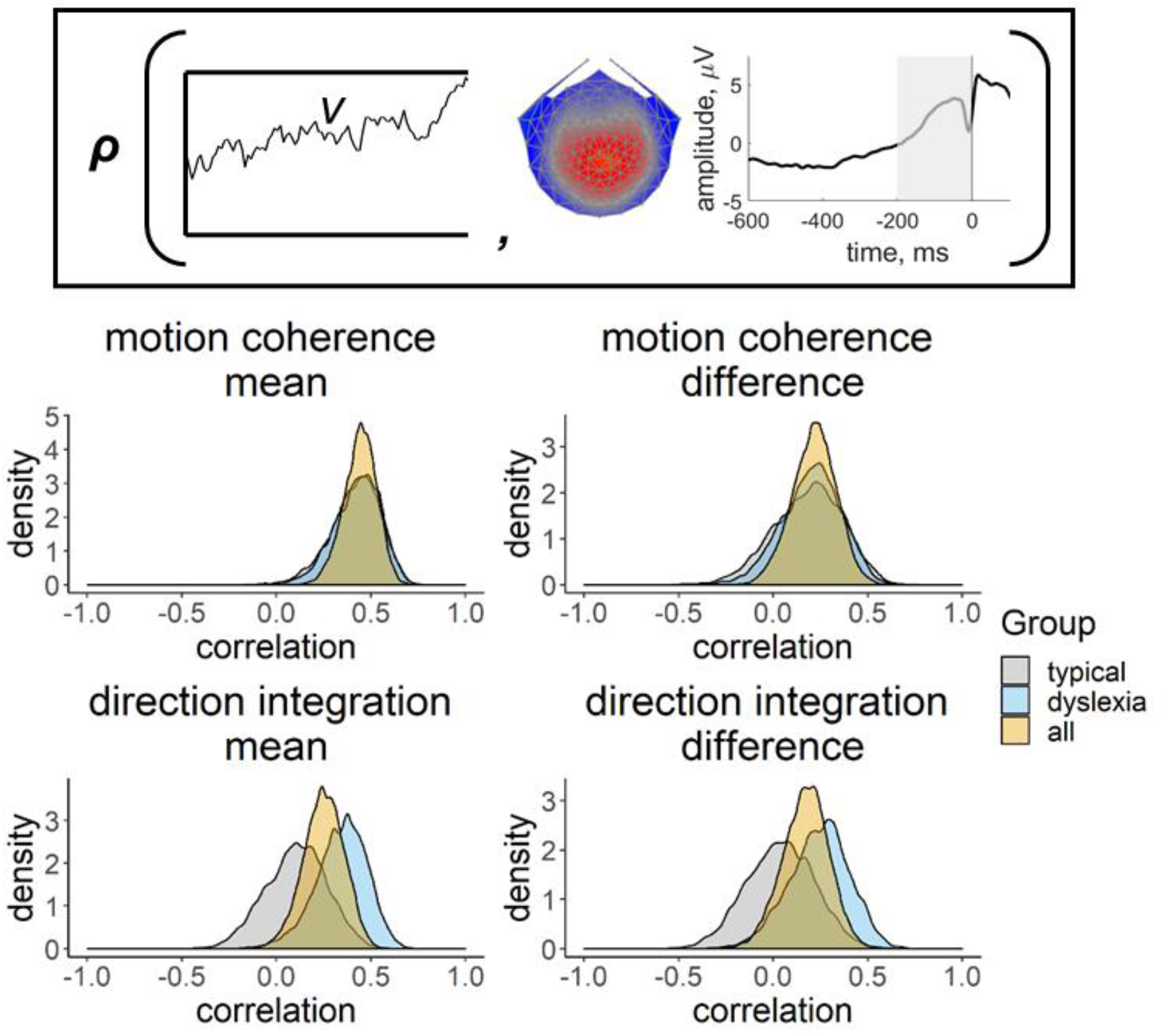
Posterior density plots showing the correlation between drift-rate and the EEG measure. Inset provides a schematic representation of the drift-rate parameter (*v*; left) and EEG measure (slope of response-locked waveform from -200 ms to 0 ms around the response; right) that were correlated in the joint model, where ***ρ*** represents the correlation. Posterior density plots in the left column reflect the correlation between the mean drift-rate across difficulty levels *(v.mean)* and the mean EEG slope measure across difficulty levels (*EEG.mean*). Posterior density plots in the right column reflect the correlation between the difference in drift-rate between difficulty levels (*v.diff*) and the difference in EEG slope measure between difficulty levels (*EEG.diff*). Plots for the motion coherence task are presented in the upper row and plots for the direction integration task are presented in the lower row. The orange distribution shows the correlation across all participants, and the grey and blue distributions show separate correlations estimated for typical children and children with dyslexia, respectively.

## Discussion

We analysed the performance of children with dyslexia and typically developing children in two global motion tasks using diffusion modelling, to identify the processing stages that are altered in dyslexia. In both the motion coherence and direction integration tasks, we found that children with dyslexia accumulated sensory evidence at a slower rate than typically developing children, once controlling for differences in age. Moreover, we found a neural correlate of this evidence accumulation process that was attenuated in dyslexia, thus linking brain and behavioural measures with a latent model parameter.

The finding of reduced evidence accumulation for children with dyslexia during the motion coherence task echoes that of a recent study by O’Brien and Yeatman (2020) and helps to explain previous reports of elevated motion coherence thresholds in dyslexic individuals (Benassi et al., 2010). Importantly, the current study goes further by showing that reduced evidence accumulation is also found in a direction integration task that does not require segregation of signal-from-noise. This result suggests that dyslexic individuals have general difficulties with extracting global motion information, rather than solely having difficulties with noise exclusion (cf. Conlon et al., 2012; Sperling et al., 2006). These general difficulties could reflect reduced temporal and/or spatial integration of motion signals (Benassi et al., 2010; Hill & Raymond, 2002; Raymond & Sorensen, 1998). This conclusion does not negate the possibility that dyslexic individuals face additional difficulties when segregating signal from noise, as we suggested based on stimulus-locked analyses using a similar dataset (Toffoli et al., under review).

By supplementing our diffusion modelling analysis with EEG, we have identified a neural index of reduced evidence accumulation in dyslexia. Specifically, we used a data-driven component decomposition technique to find a centro-parietal component resembling that previously linked to the decision-making process (Kelly and O’Connell, 2013; O’Connell et al., 2012; Manning et al., 2021), and then used a linear deconvolution technique to ‘unmix’ overlapping stimulus- and response-locked activity. We found that children with dyslexia showed a more gradual build-up in the response-locked centro-parietal component compared to typically developing children, and the gradient of the build-up was positively correlated with drift-rate in the joint model. While the EEG analysis was exploratory, with the precise analysis steps not pre-registered, the results are consistent with an earlier study of typically developing children which linked build-up in a comparable centro-parietal component with drift-rate (Manning et al., 2021) and follow the pattern we hypothesised (https://osf.io/enkwm).

Alongside reductions in drift-rate, we hypothesised that children with dyslexia would show wider boundary separation compared to typically developing children, reflecting more cautious responses, and no differences in non-decision time. We found some evidence for increased boundary separation in children with dyslexia in the motion coherence task, but this was inconclusive evidence. There was also little evidence for group differences in non-decision time, but the evidence was again inconclusive. These inconclusive results are therefore not at odds with O’Brien and Yeatman (2020), but suggest that more data would be required to reach a firm conclusion regarding these parameters. Therefore it seems that any group differences in these parameters are more subtle than group differences in drift-rate. It is worth noting that the inferential method used by O’Brien and Yeatman (2020) differed from our own: while they also fit a hierarchical Bayesian model, they then extracted point estimates of diffusion model parameters for each individual which they used to draw statistical inferences. Importantly, this means that the approach of O’Brien and Yeatman (2020) ignored the uncertainty in the individual-level parameters, which can inflate the evidence in favour of the winning model (Boehm et al., 2018; Evans & Wagenmakers, 2019).

Together with the results from stimulus-locked analyses using the same dataset (Toffoli et al., 2020), our results suggest that early sensory encoding of motion information is not altered in children with dyslexia. In the current study we found no evidence of group differences in non-decision time – a measure which includes the time taken for sensory encoding, and Toffoli et al. showed that early peaks reflecting motion-specific processing were similar in children with dyslexia and typically developing children, with differences arising only after ∼430 ms following stimulus onset, specifically in the motion coherence task. The current analyses show that differences in dyslexia arise due to the efficiency with which evidence is extracted from global motion stimuli and integrated towards a decision bound, which is often attributed to parietal areas (Hanks et al., 2006; Shadlen & Newsome, 1996; 2001; de Lafuente et al., 2015). Without a comparable form task, it is unclear from the current study whether reduced evidence accumulation is restricted to tasks that rely heavily on the dorsal stream. However, we suggest that *within* the magnocellular/dorsal stream, early sensory processing is unaffected in dyslexia (cf. Livingstone et al., 1991; Stein, 2001, 2019; Stein & Walsh, 1997), with group differences emerging only at later processing stages, including those involved in decision-making. Future work will be required to determine how specific reduced evidence accumulation in dyslexia is to visual motion processing. Slower responses have been reported in dyslexia for other tasks (Catts et al., 2002, Nicolson & Fawcett, 1994) which could reflect pervasive reduced evidence accumulation. However, slowed responses could arise for different reasons (e.g., increased non-decision time, or wider boundary separation), so diffusion model decompositions on a range of tasks are required.

A number of future directions for further research in this area emerge. What cognitive skills other than magnocellular / dorsal stream processing contribute to reduced drift-rate in dyslexia? As highlighted by O’Brien & Yeatman (2020), future research will need to assess whether and how reduced evidence accumulation is related to rapid automatized naming (RAN) skills or processing speed measures more generally. General processing speed is a unique predictor of word reading and comprehension (Christopher et al., 2012) and RAN is a recognized independent contributor to variation in reading ability, complementing phonological skills (e.g., O’Brien & Yeatman, 2020). Future work will need to establish the extent to which reduced processing speed and slower RAN associate with reduced drift-rate in dyslexia. In addition, performance IQ varied across our two samples and was associated with drift-rate. Exploratory models revealed that, even when controlling for both age and performance IQ, there was still relatively more evidence for group differences in drift-rate than for a lack of group differences. Yet the strength of evidence was weaker than in models controlling only for age. Importantly, partialling out differences in performance IQ could lead to removing some of the variance related to the group differences we are interested in, as atypical development could lead to both dyslexia and reduced IQ (Dennis et al., 2009). Indeed, performance IQ has been shown to strongly predict reading skills, independently of phonological skills (O’Brien & Yeatman, 2020). Future work will need to investigate the contribution of processing speed and performance IQ to decision making in dyslexia as a whole spectrum, for subgroups of children with reading difficulty (Bonifacci & Snowling, 2008), and across typical development more generally.

By combining diffusion modelling and EEG measures that are sensitive to the multiple processes contributing to visual motion perception, we have uncovered differences between children with dyslexia and typically developing children that could not be observed in children’s behavioural responses alone. Moreover, diffusion modelling provides a way to measure sensitivity to motion information without confounding speed-accuracy tradeoffs. Given that reduced behavioural sensitivity to motion has been reported in a range of developmental and psychiatric conditions (Braddick et al., 2003; Chen et al., 2003; McKendrick & Badcock, 2004), we suggest that diffusion modelling may provide a useful framework to identify whether similar or distinct processing stages are affected in different conditions, with implications for understanding the development of these conditions and their relationship to other cognitive processes.

## Data Availability

Data will be made available on the UK Data Service after the manuscript has been accepted for publication.

https://osf.io/nvwf7

## Acknowledgements

We are grateful to the participants and families who took part, the schools and organisations who kindly advertised the study, Irina Lepadatu, the Oxford Babylab and Dhea Bengardi for help with recruitment, and Helena Wood, Lisa Toffoli, Madeleine Mills, Amber Heaton, and Kate Seaborne who helped with data collection and data entry. The project was funded by a Sir Henry Wellcome Postdoctoral Fellowship awarded to CM (grant number 204685/Z/16/Z) and a James S. McDonnell Foundation Understanding Human Cognition Scholar Award to GS. NJE was supported by an Australian Research Council Discovery Early Career Researcher Award (DE200101130). We are grateful to Dorothy Bishop for providing funding for research assistance.

## Supplementary Materials

**Figure S1.**
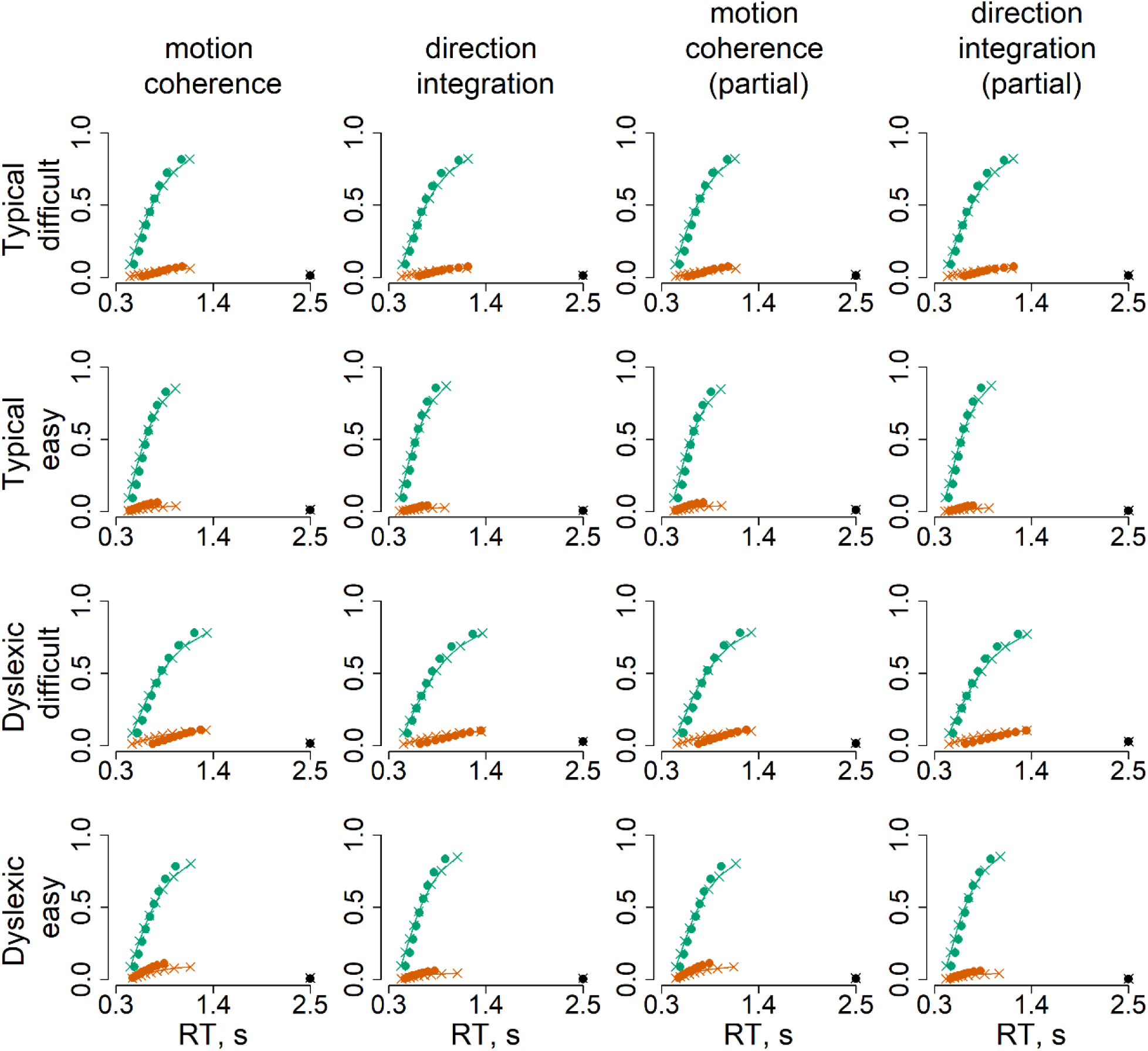
Model fits. Defective cumulative density function plots for each of the four models, for typically developing children (upper rows) and children with dyslexia (bottom rows) for difficult and easy levels. Green represents correct responses and red represents error responses, at each of 9 quantiles. The dots reflect the observed data and crosses with connecting lines reflect the model fit. The dots and crosses at 2.5 seconds reflect the observed and model predicted misses.

**Figure S2.**
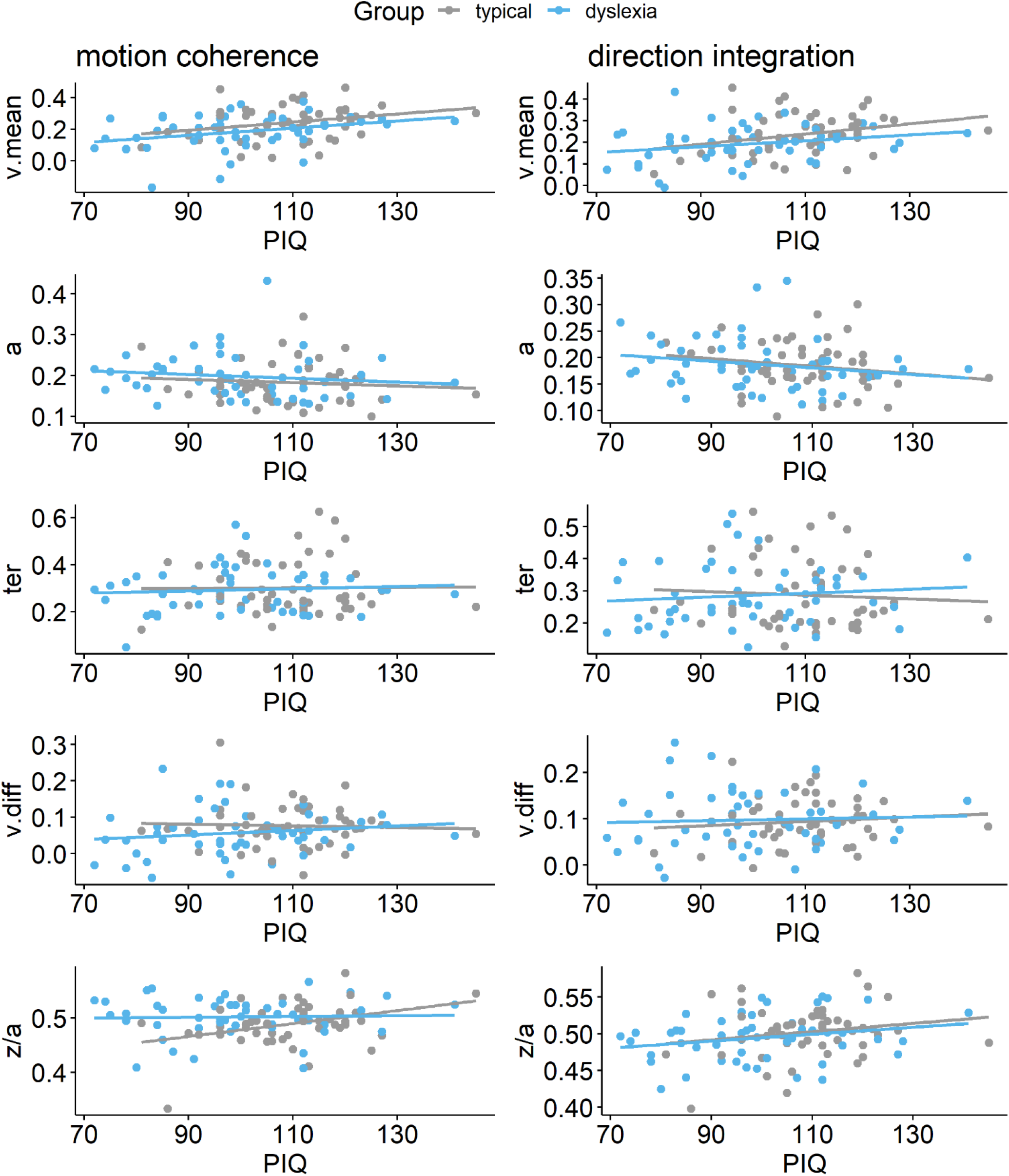
Scatterplots plotting individual parameter estimates against performance IQ. Maximum likelihood estimates contained within the posterior for each participant’s mean drift-rate across difficulty levels (*v.mean*), boundary separation (*a)*, non-decision time (*ter*), difference in drift-rate between difficulty levels (*v.diff*), and starting point *(z/a)*, plotted as a function of performance IQ (PIQ), for the motion coherence task (left column) and direction integration task (right column). Typically developing children are plotted in grey and children with dyslexia are plotted in blue.

**Figure S3.**
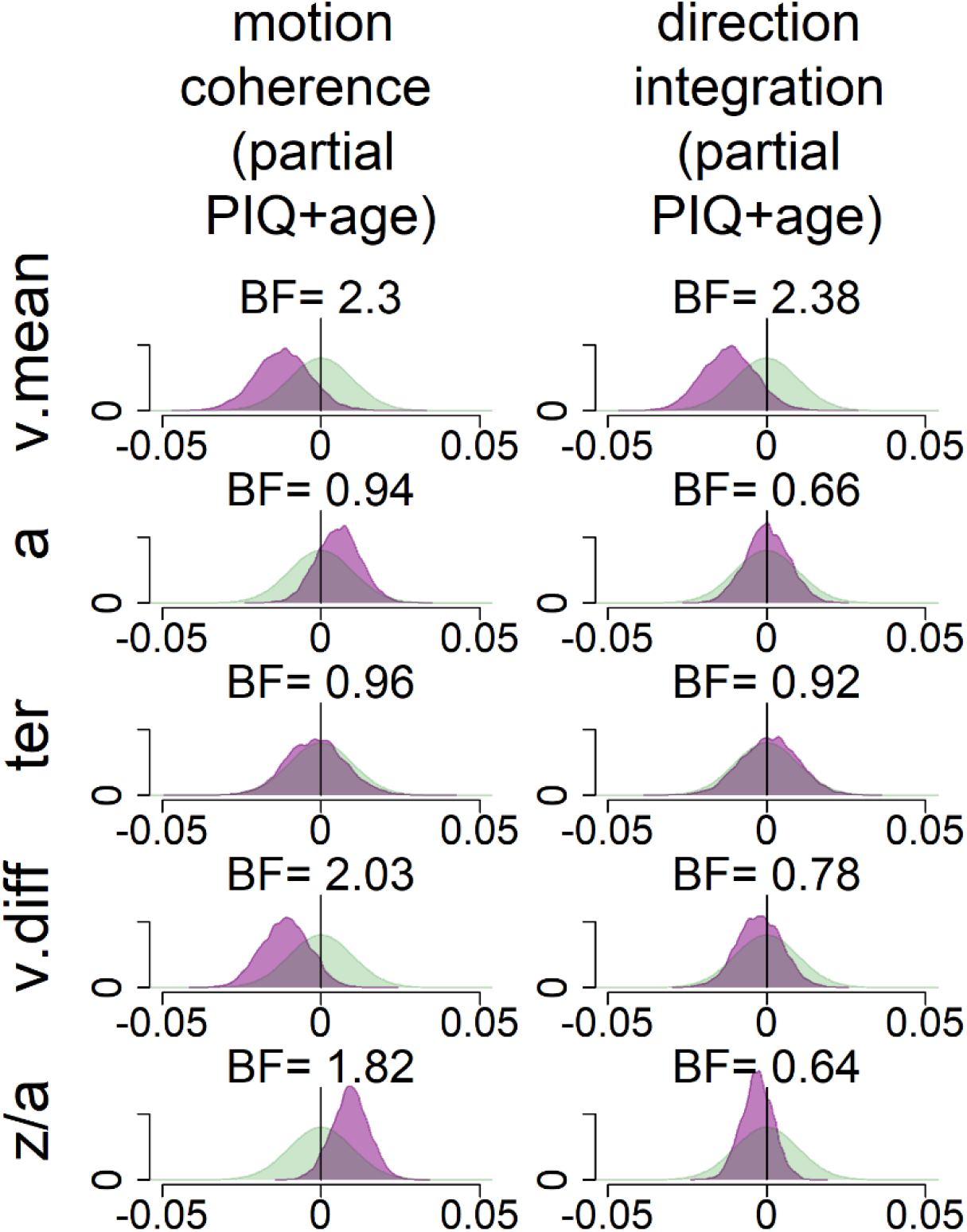
Exploratory analyses: prior and posterior density distributions for model with age and performance IQ partialled out. While our pre-registered analysis did not control for performance IQ, we conducted an exploratory analysis to investigate whether group differences in drift-rate were still apparent when controlling for performance IQ. The figure shows prior (green) and posterior (purple) density distributions for the group-level parameters reflecting group differences in each of the 5 model parameters (v.mean = mean drift-rate across difficulty levels; a = boundary separation; ter = non-decision time; v.diff = difference in mean drift-rate between difficulty levels; z/a = relative starting point) for each task, when both age, performance IQ (PIQ) and their interaction are partialled out. Negative values reflect lower parameter values in the dyslexia group compared to the typically developing group. BF = Savage-Dickey Bayes factors in favour of the alternative hypothesis (H_1_) over the null hypothesis (H_0_). BF > 1 support H_1_. As in Figure 7, the posterior distribution for *v.mean* is shifted leftwards, reflecting lower mean drift-rate in the dyslexia group than the typically developing group. The corresponding Bayes factors are smaller in these analyses, indicating weaker evidence for group differences. As we reflect on in the Discussion of the main manuscript, the decision to partial out PIQ should not be taken lightly, as PIQ seems to contribute to both decision making variables (drift-rate) and group differences, so it is likely that partialling out PIQ removes some of the variance related to the group differences we are interested in.

**Figure S4.**
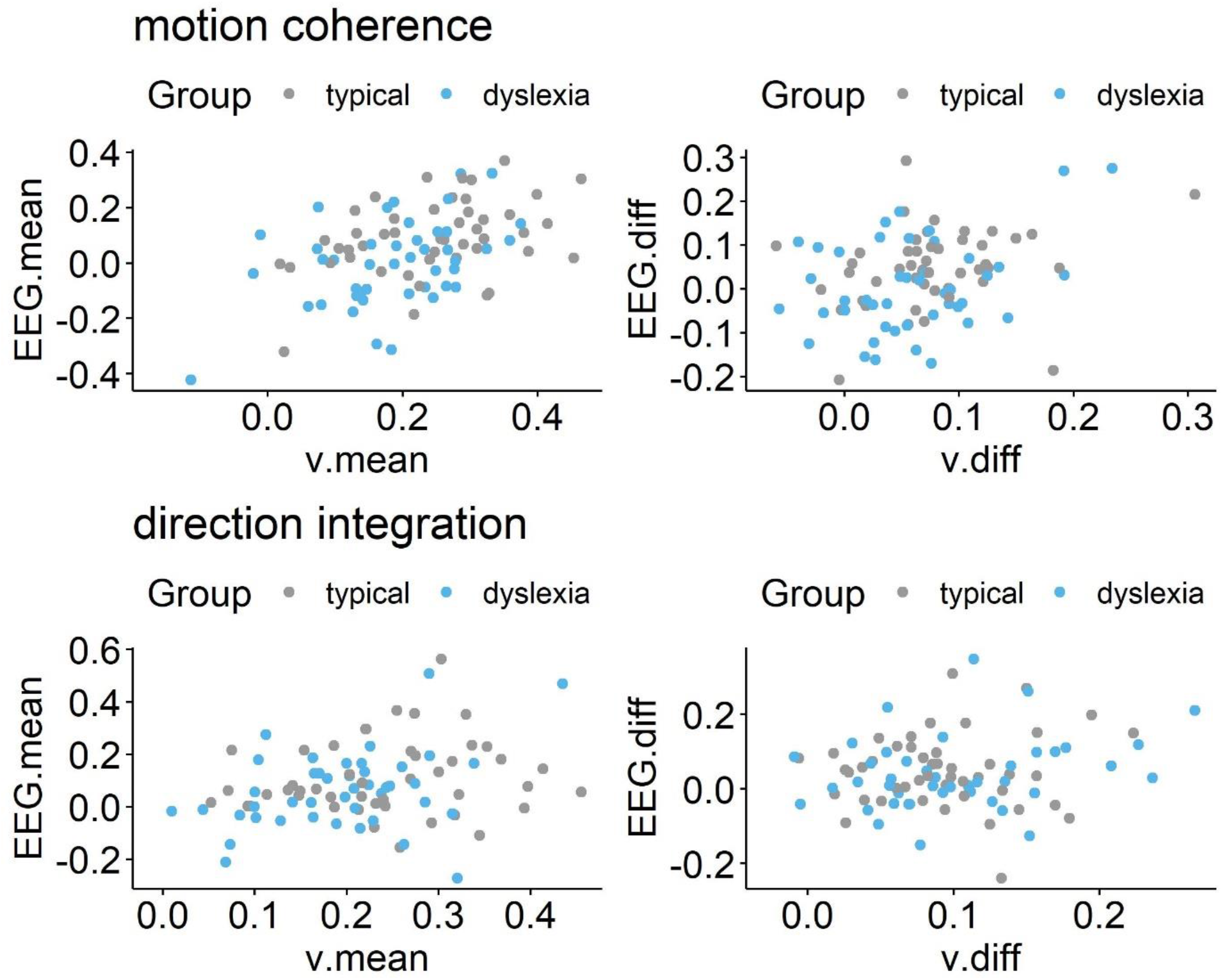
Scatterplots showing relationship between drift-rate and EEG. Left panels show maximum likelihood estimates contained within the posterior for each participant’s mean drift-rate across difficulty levels (*v.mean*) plotted against the slope of EEG activity averaged across difficulty levels (*EEG.mean*) for the motion coherence (top) and direction integration (bottom) tasks. Right panels show point estimates for each participant’s difference in drift-rate between difficulty levels (*v.diff*) plotted against the difference in slopes of EEG activity between the two difficulty levels (*EEG.diff*), for each task. Typically developing children are plotted in grey and children with dyslexia are plotted in blue.

